# Disability Training for Health Workers: A Global Narrative Systematic Review

**DOI:** 10.1101/2021.08.03.21261522

**Authors:** Sara Rotenberg, Rodríguez Danae Gatta, Azizia Wahedi, Rachelle Loo, Emily McFadden, Sara Ryan

## Abstract

**Background:** Health worker training on disability is a recognized component of achieving high standards of health for people with disabilities, given that health worker’s lack of knowledge, stigma, and negative attitudes towards people with disabilities act as barriers to high quality health care.

**Objective:** To understand the published literature on training health workers about disability.

**Methods:** We searched five databases for relevant peer-reviewed articles published between January 2012 and January 2021. Studies that focused on training health care workers to improve knowledge, confidence, self-efficacy, and competence to support people with physical, sensory, or intellectual impairments were included. Data about the details of the intervention (setting, participants, format, impact assessments, etc.) and its effects were extracted.

**Results:** There is an array of highly local tools to train health workers across stages of their training and careers (pre-service, in-service, and continuing professional development). Studies involving people with disabilities in the training, community placements, simulations, or interactive sessions were found to be most effective in improving knowledge, confidence, competency, and self-efficacy.

**Conclusions:** As part of initiatives to build inclusive health systems and improve health outcomes for people with disabilities, health workers around the world need to receive appropriate and evidence-based training that combine multiple methods and involve people with disabilities.

## BACKGROUND

Human resources for health are at the heart of high-quality health systems. It is critical to improve health worker training to improve health care for populations that are systematically marginalized by health systems, such as people with disabilities. The World Health Organization (WHO) estimates that people with disabilities make up 15% of the world’s population.^1^ People with disabilities often face significant barriers to health care, including lack of accessible transport and facilities, limited financial protection, poor health worker attitudes that result in worse outcomes or limited health worker training on disability.^2^ Even in countries where there is guaranteed universal access and financial protection, health workers’ unfamiliarity with disability, or negative attitudes towards people with disabilities, can not only foster an unwelcoming environment, but also contribute to high rates of patient safety issues and poor quality care.^1^

Health worker training on disability is a recognized component of achieving high standards of health for people with disabilities. While the UN Convention on the Rights of Persons with Disabilities (UNCRPD) Article 25^3^ has specific requirements on access to health care and SDG monitoring on health worker disability training, the recent World Health Assembly resolution most eminently highlights the role of health worker training in removing barriers to health care for people with disabilities.^4^ In addition, recent studies have highlighted the need to improve health care workers’ attitudes, knowledge, and competency to provide care for people with disabilities. For example, a US study illustrated that just 40.7% of physicians were confident about providing care to patients with disabilities and most (82.4%) perceived that people with significant disabilities have worse quality of life. Similarly, a study found that 87% of nursing students implicitly associated negative traits with physical disability, which may influence clinician behaviour. These studies illustrate the need to improve health workers’ confidence, competency, attitudes, and comfort in treating patients with disabilities. Given these international agreements and recent studies, it is important that countries around the world begin to integrate disability training systematically and use examples of successful interventions as models.

This review directly builds on a previous review by Shakespeare and Kleine that explored health worker training on disability between 2000 and 2011.^5^ The study found that, while there are numerous interventions to teach medical professionals about disability, there are few common philosophical underpinnings, insufficient hands-on experience, and more opportunities to incorporate disability across the curriculum.^5^ Since this review, additional systematic reviews have examined health worker training on people with disabilities for certain populations of health workers^6^, certain impairments,^7^ or geographic areas.

Given renewed international commitments to health worker training on disability and country-level plans in Australia^8^ and the UK^9^ to train health workers on specific types of disability, it is important to update Shakespeare and Kleine’s review^5^ and outline the types of interventions to improve health worker’s knowledge, confidence, self-efficacy, and competence in treating patients with disabilities. Understanding the ways in which health workers receive training on disability, will facilitate adjusting system-level policies and individual-level practices to improve care for people with disabilities. Ultimately, this review will help to understand the types of training that support positive and sustained improvements in service delivery for health workers serving people with disabilities.

## METHODS

### Search Strategy

Electronic searches were conducted for the EMBASE, Global Health, Medline, CINAHL, ASSIA and Web of Science databases between 18-19 January 2021. Search terms were developed in three domains: disability, health education, and health workers. Disability terms were general, focusing on various types of impairments; health education terms targeted aspects of health training (i.e., ‘core competency’, ‘patient encounter’, ‘standardized patient’, etc.); and health worker terms were developed using key terms from WHO’s International Classifications of Health Workers.^10^ Terms were developed using MeSH, keywords, or equivalent as well as from other reviews on similar topics and searches were limited to papers in English, French, or Spanish. These parameters and strategy were agreed upon by the authors and a research librarian before the search was conducted to ensure there were adequate words to capture articles across the three domains examined. The Preferred Reporting Items for Systematic Reviews and Meta-Analysis (PRISMA) statement was followed for conducting and reporting the review (PROSPERO Registration: CRD42021231120). All studies identified by the review were exported into an EndNote database (version X20, Clarivate Analytics, Philadelphia, PA, USA) and then exported into Rayyan (Qatar Computing Research Institute, Qatar) for screening. ^11^

### Selection Criteria

Our search strategy sought to identify peer-reviewed articles from around the world published between January 2012 and 2021. Given a previous systematic review covered this topic until 2011^5^, the search included articles published from 2012-January 2021 and included all health worker types, health education levels, and disability globally. The inclusion criteria required that studies were: qualitative and/or quantitative in methods; included a complete description of the intervention; explicit evaluation of the training’s impact (i.e., pre- and/or post-training evaluations, follow-up surveys, etc.); and had a particular focus on improving disability competency, knowledge, confidence, self-efficacy, curricula, or teaching methods. Studies examining health worker attitudes towards people with disabilities were excluded on the basis that a positive attitude does not necessarily guarantee improved competency or care outcomes. Studies that measured attitudes alongside other criteria were included. Finally, the abundance of articles on training health workers about mental health, the authors decided that this topic merited further, independent exploration, and, therefore, we excluded papers that trained health workers only about mental health. Only papers that looked at physical, sensory, intellectual or developmental impairments were included.

### Data Extraction

All data were extracted into a Google Sheet developed for this review. 78 full-text articles underwent data extraction, following a title, abstract, and full-text review by two reviewers (SRo and DR). An additional three reviewers (SRo, AW, RL) extracted data related to the general study information, setting, country, health worker cadre, number of participants, type of disability, features of the intervention, impact measurement, and outcomes. The extraction was double-checked by a second reviewer and collectively checked again by the extractors (SRo, AW, RL). Any conflicts in inclusion or extraction were resolved through discussion with a third and/or fourth member of the review team.

Given the wide array of study instruments and outcomes used to assess training impact, a meta-analysis could not be conducted, and a narrative synthesis was conducted instead. Quality and bias assessments were conducted by at least two reviewers in accordance with the SIGN50 (Scottish Intercollegiate Guidelines Network Checklists) and modified slightly from previous methods.^13^

Studies were rated as low bias, if all or almost all of the criteria were fulfilled, and those that were not fulfilled were thought unlikely to alter the conclusions of the study; medium, if some of the criteria were fulfilled, and those not fulfilled were thought unlikely to alter the conclusions of the study; or high, if few or no criteria were fulfilled, and the conclusions of the study were thought likely or very likely to alter with their inclusion.

## RESULTS

The preliminary search identified 5,665 articles for title and abstract screening, after 1,527 duplicates were removed. Following screening, 247 articles were included for full-text review. Twelve studies were excluded because full-texts could not be retrieved and a further 154 studies did not meet the inclusion criteria as shown in Fig.1., and 3 articles included in the review were excluded during extraction because of an unclear intervention (n=1) and wrong population group (n=2).^14^

**Fig 1.**
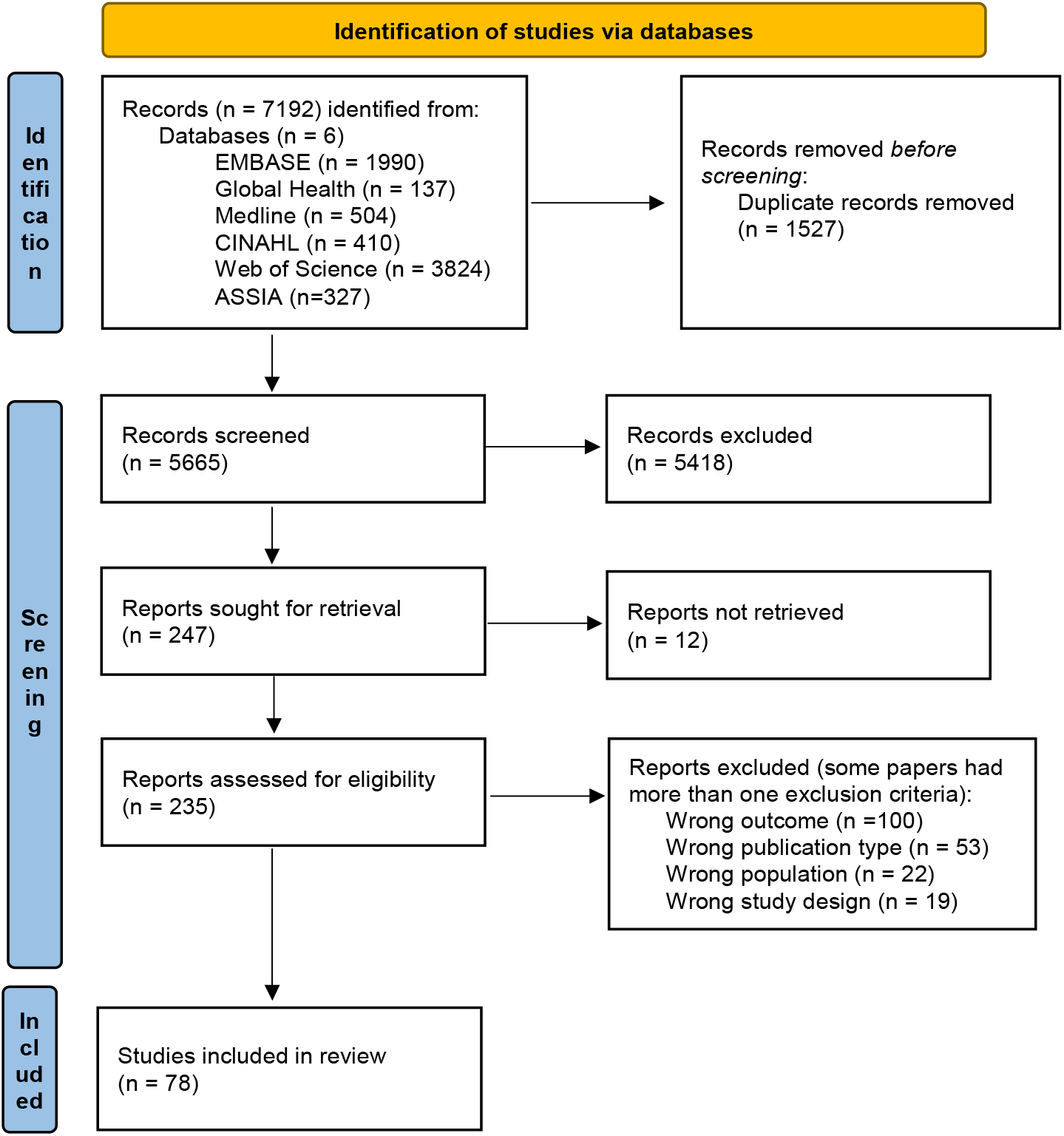
Flow chart of selected studies to review health worker training on disability ^12^.

Included studies (n=78) represented a range of geographies, health workers, and intervention types. Among these, there were studies from 19 countries, including seven low- and middle-income countries.^15^ Most studies took place in the United States (n=35), followed by the United Kingdom (n=13). Among the studies included, 30 were rated as low, 43 as medium, and five as high risk of bias [Supplementary Materials]. Studies varied in whether they were mandatory or optional; free or paid; and for certification or elective; however, many studies did not include this information. Various cadres of health workers were included in the study; doctors, medical students or residents (n=37), and nurses or nursing students (n=17) were the main recipients of training. These health workers were generally trained in the pre-qualification stage (n=52), though there were several in-service (n=7) and continuing professional development (CPD) programs (n=19). The review included studies across disability groups; the most common focus was training about people with intellectual and developmental disabilities (n=41), followed by general programs about people with disabilities (n=16). Most studies measured improvements in knowledge (n=57) and competence (n=42) outcomes, yet most studies used a self-designed evaluation instrument (n=54). There was a wide variety of techniques to train health workers about disability, including lectures or other didactic methods (n=65), and case studies (n=28); the majority of studies (n=58) used multiple teaching modalities [Appendix A].

### Lecture/Didactic Methods

Most (n=65) studies included lectures or didactic methods, such as videos, multi-media formats, or online coursework. Many studies used these opportunities to introduce health professionals to the topic of disability from a rights-based perspective to enhance attitudes, awareness, and knowledge about disability. Some studies also taught particular skills that could be applied in-practice, such as an elective sign language class for medical and pharmacology terms^16^ to improve skills for engaging with d/Deaf or hard of hearing individuals. Lectures were often combined with some other intervention (case study or simulation) to apply knowledge learned from the lecture. Participants in combined programs identified the content to be quite engaging and contributed to greater improvements in key outcomes. However, for those who only completed lecture or didactic-based methods, there were still improvements in the general outcomes, but it was often less significant than studies that combined multiple methods ^14,17,18.^Finally, some programs utilized novel, innovative technology and multimedia tools to teach about disability in a more engaging method than traditional lectures or didactic methods. For example, one study designed multi-media tools (MMLTs) to teach medical students about common visual impairments and compared the knowledge scores with those who had read a textbook. The findings highlight the importance of engaging material, as while there was no significant difference in knowledge (except for cataract recognition), the MMLT took less time and 87% of individuals found it more enjoyable than traditional teaching methods.^19^

### People with Disabilities as Teachers

Recognizing the important role of self-advocates and patients as educators^5^, several studies (n=19) invited people with disabilities to share their experiences in the health system, portray standardized patients, or give a lecture. Some universities hired people with disabilities to participate in simulated patient programs, while others asked patients to participate voluntarily. Many others found creative ways of engaging with people with disabilities. Cardiff University, for example, hired a self-advocacy theatre group to run a simulation and icebreaker activity.^20^ These activities added a non-clinical dimension to medical training about disability, as it allowed participants to explore disability outside the health worker-patient relationship and engage in dialogue. Studies that measured participants comfort and attitudes before and after a person with a disability as a teacher demonstrated that participants felt the non-clinical interaction enhanced their comfort and attitudes towards people with disabilities. ^21, 22^

**Table 1:** Characteristics of Included Studies.

| Variable | Number | Percentage (%) |
| --- | --- | --- |
| **Training Location** |  |  |
| North America | 40 | 51.2 |
| Europe | 22 | 28.2 |
| Asia | 6 | 7.7 |
| Oceania | 4 | 5.1 |
| Africa | 4 | 5.1 |
| South America | 2 | 2.5 |
| **Disability Focus of Training\*\*†** |  |  |
| Intellectual and developmental | 41 | 51.9 |
| All types of disabilities/unspecified | 16 | 20.3 |
| Physical/motor | 13 | 15.2 |
| Communication | 9 | 11.4 |
| D/deaf and hard of hearing | 8 | 10.1 |
| Visual impairment | 3 | 3.8 |
| Sensory impairment | 2 | 2.5 |
| **Health Worker Type\*\*†** |  |  |
| Medics | 37 | 35.2 |
| Nurses | 17 | 16.2 |
| Occupational and Physical Therapists | 17 | 15.2 |
| Allied Health Professionals | 8 | 7.6 |
| Dentists | 7 | 6.7 |
| Audiologists and Speech Language Pathologists | 6 | 5.7 |
| Psychologists | 4 | 3.8 |
| Personal Care Workers | 5 | 4.8 |
| Community Health Workers | 2 | 1.9 |
| Pharmacists | 2 | 1.9 |
| **Teaching Method\*** |  |  |
| Lecture/didactic methods | 65 | 34.7 |
| Case study | 28 | 14.9 |
| Clinical encounter | 26 | 13.9 |
| Placements, experiential, and community-based learning | 25 | 13.4 |
| Simulation | 24 | 12.8 |
| People with disabilities as a teacher | 19 | 10.1 |
| **Intervention Outcomes\*** |  |  |
| Knowledge | 57 | 28.4 |
| Competence | 42 | 20.9 |
| Attitudes | 31 | 15.4 |
| Confidence | 24 | 11.9 |
| Comfort | 15 | 7.5 |
| Communication skills | 12 | 6.0 |
| Self-Efficacy | 11 | 5.5 |
| Other related outcomes | 9 | 4.5 |
| *Total number is greater than 78 because studies included multiple variables |  |  |
| †categories include students and residents who are in training to become fully qualified as this type of health worker |  |  |

### Case Studies

Case studies are a common tool in medical education to prepare health workers holistically for their education, provided they have clear structure, details, and observations.^23^ Accordingly, many studies used case studies (n=28) as a way of learning how to improve care for people with disabilities. These tools were especially common in continuing professional development, as some programs ask patients to bring case examples to review anonymously to improve care ^24, 25^ or were to spur reflection on their own work. Several case studies were conducted through online learning or innovative interactive methods. For example, the City University of London created CitySCaPE, which is a multi-media simulation that simulated different patient cases of people with intellectual disabilities with nursing students. ^26^ These types of case studies that blended the traditional case study and simulation aspects created greater engagement in settings where in-person or clinical encounters were not possible.

### Placements, experiential, and community-based learning

Placements, experiential, and community-based learning (n=25) methods were sustained opportunities to engage with people with disabilities in clinical and alternative settings that were common in in-service and pre-qualification training. For example, some studies examined the impact of clerkship placements in specialized clinics for people with disabilities,^27^ while others looked at nurses and occupational therapists’ improvements after participating in a week-long summer camp for children with disabilities.^28^ In the clinical setting, students found that they improved skills because they were able to engage with people with disabilities for extended periods, rather than a singular interaction. Furthermore, the out-of-clinic engagement, such as at camps, schools, or residential settings helped illustrate the non-medical and everyday lives of people with disabilities.

### Simulations

Simulations were found to be helpful tools to support skill development and learning outside of health worker-patient interactions. Many speech-language pathology, nursing, and medical student programs used simulations (n=24) as tools to develop confidence and communication skills when treating patients with communication disorders.^29,30, 31^ In addition, several medical schools integrated disability training into existing clinical simulation skills labs to improve care for people with disabilities. For example, at the University of Gothenburg medical students were videotaped during a simulated patient exercise to reflect on improving communications skills, particularly for the simulated patient with an acquired communication disorder.^31^ Overall, the simulations were useful tools for improving knowledge, comfort, and competency in a low-pressure environment that is applicable to serving people with disabilities.

### Clinical Encounters

Several programs (n=26) included singular clinical encounters with patients with disabilities as part of their disability training. These were often day-long programs to familiarize students with providing care and were predominantly focused on improving knowledge and competency.^32^ Most students who participated in these programs were advanced (i.e., penultimate or final year students) who had previously had some education on providing care to people with disabilities. These opportunities focused on practicing clinical skills to treat patients with disabilities, and, despite the short exposure, did significantly affect participant’s key outcome scores. For instance, clinical encounters used in CPD, such as in Rwanda, where instructors in a physiotherapist training program went to participants’ clinics to provide immediate feedback on their practice.^33^

### Multi-pronged approach

Approximately 75% of papers utilized a combination of methods to have impact on training participants. These multi-pronged approaches helped reach various learning styles and cement learning. Two papers included in this review utilized all of the interventions measured in this paper. For example, two State University of New York medical colleges demonstrated the importance of integrating disability across health worker curricula, as all participants significantly improved their knowledge, attitudes, and core competencies in treating patients with disabilities.^34^ Similarly, the University of South Florida had a 12-week clinical clerkship that involved classroom simulations, lectures, case studies, people with disabilities as teachers, and a twice-weekly placement in a community clinic that served people with disabilities. The immersion helped to significantly improve knowledge, attitudes, and comfort.^35^

## DISCUSSION

Numerous studies and examples serve as successful models to train health workers about disability and improve knowledge, competence, skills, self-efficacy, and confidence to treat patients with disabilities. All teaching methods had some positive impact on the outcomes measured in this study, regardless of health worker type, location, or training stage, though the most commonly used were lecture/didactic methods and case studies. Part of the success of these programs was the multi-pronged nature of the approach, as 75% of studies used multiple teaching methods. The two examples that utilized all teaching interventions demonstrate the importance of a multi-pronged approach that emphasizes mainstreaming disability in health curricula, either through sustained engagement in a curricula ^34^ or an intensive placement.^35^

However, limited information about commonalities in curricula could be extracted from the data, given the diversity of interventions methods and topics.

It is important to note that these findings are not substantially different from the 2011 review.^5^ Similar methods are still used to teach health workers on disability and each example is highly localized, within either a certain school or region, other than two studies that examined national-level training programs.^33, 36^ The limited evidence of systemic integration of disability training within health worker practices is concerning in this context as the current status of training appears to depend on where you received your training, where you live, or where you work. Enacting systemic-level change to ensure all health workers have the same level of high-quality training on disability will contribute to providing consistent, high-quality care and outcomes for people with disabilities.

Similarly, there was limited standardization in tools used to measure the impact of disability training on health workers. Of the 78 studies, nearly 70% designed their own instruments, and only two studies included the same standardized measure of outcomes.^37,38^ Few studies measured the longevity of the intervention’s impact. Sustained approaches that mainstream disability should help to ensure learning is not performative for post-intervention evaluation, but actually effect change. This finding was previously noted ^5^ and there has been little improvement. Developing a common, standardized cross-disability tool or protocol for evaluating immediate and long-term intervention impact may help support monitoring and evaluation efforts to further refine and improve training.

While the review uncovered many examples of disability training, one of the main limitations of the study is that it only highlights published examples of studies, which can leave out unpublished examples. Furthermore, without greater follow-up evaluation or standardization in evaluation, it is difficult to assess the longevity and quality of impact to understand impact of training definitively. On the other hand, this study reveals some adaptable examples of how to integrate disability training into all stages of health worker training and development, which can serve as models for inclusion efforts around the world.

## CONCLUSION

Significant health disparities and poor-quality health care still exist for people with disabilities around the world. Without normalizing disability training as part of high-quality health worker training, there will continue to be limited progress on improving outcomes for people with disabilities. Catalyzing the post-pandemic health systems strengthening efforts to include these evidence-based and effective health worker training on disability can contribute to improved care and outcomes for people with disabilities.

## Data Availability

N/A

## Appendix A: Details of interventions in included studies

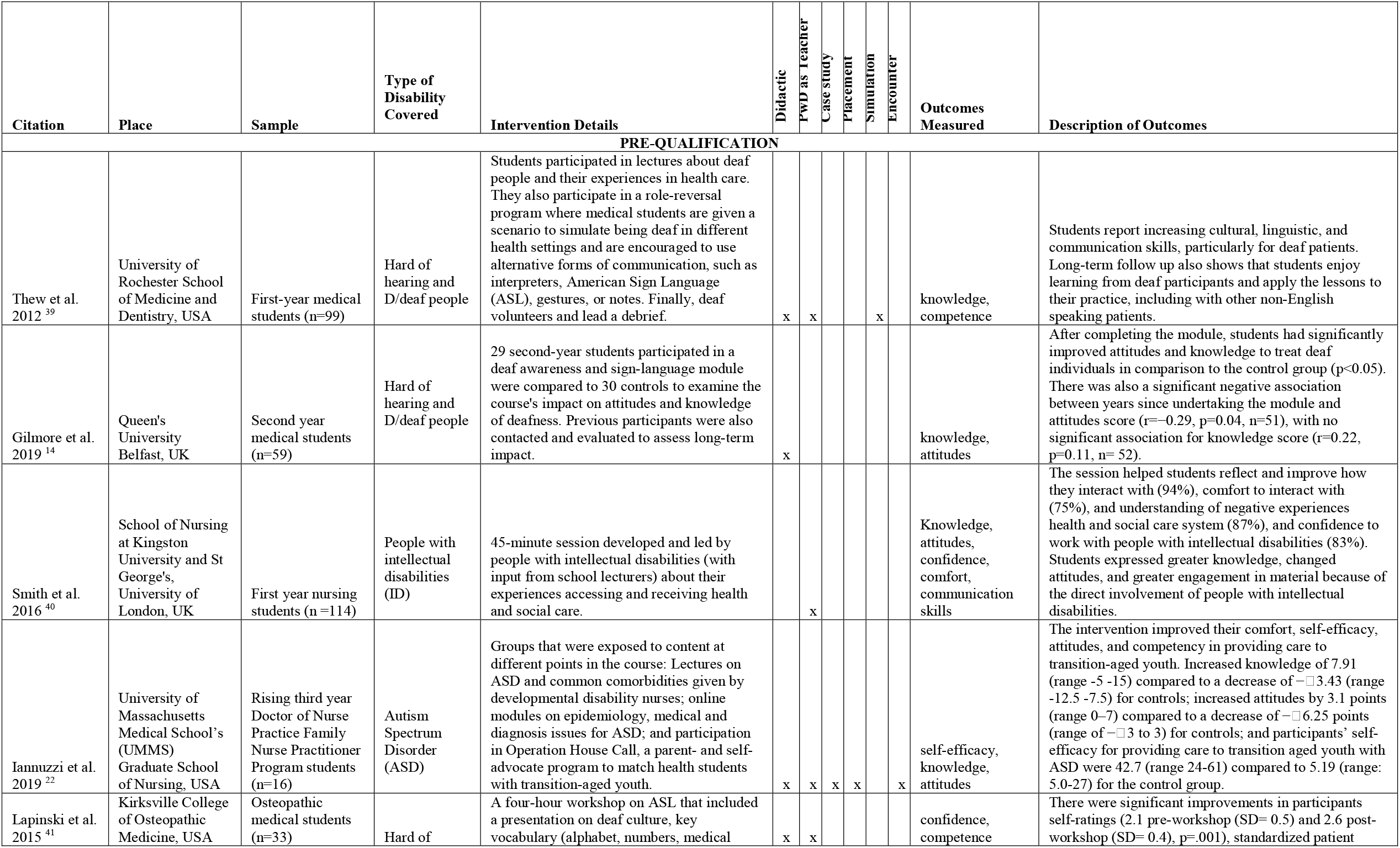
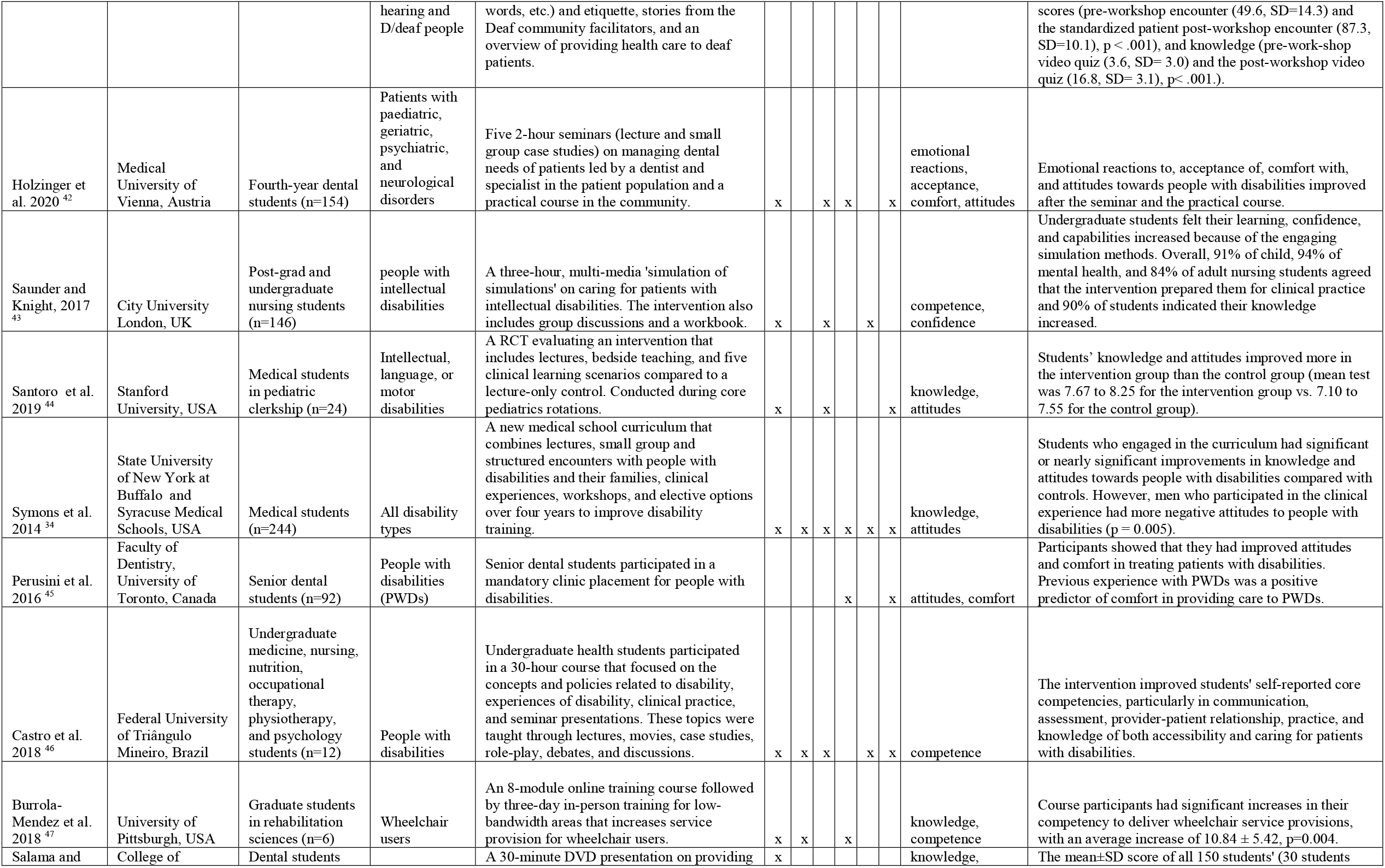
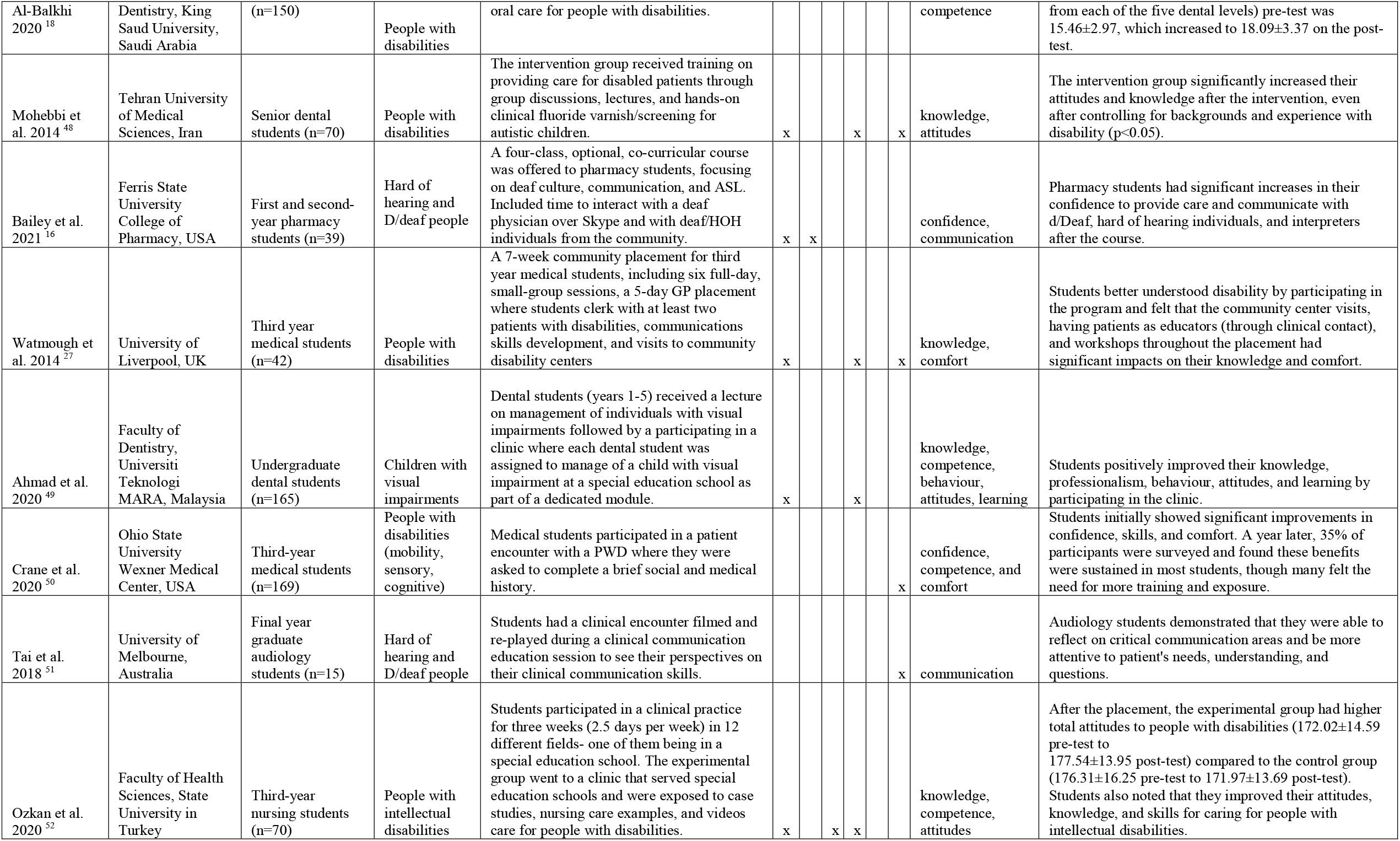
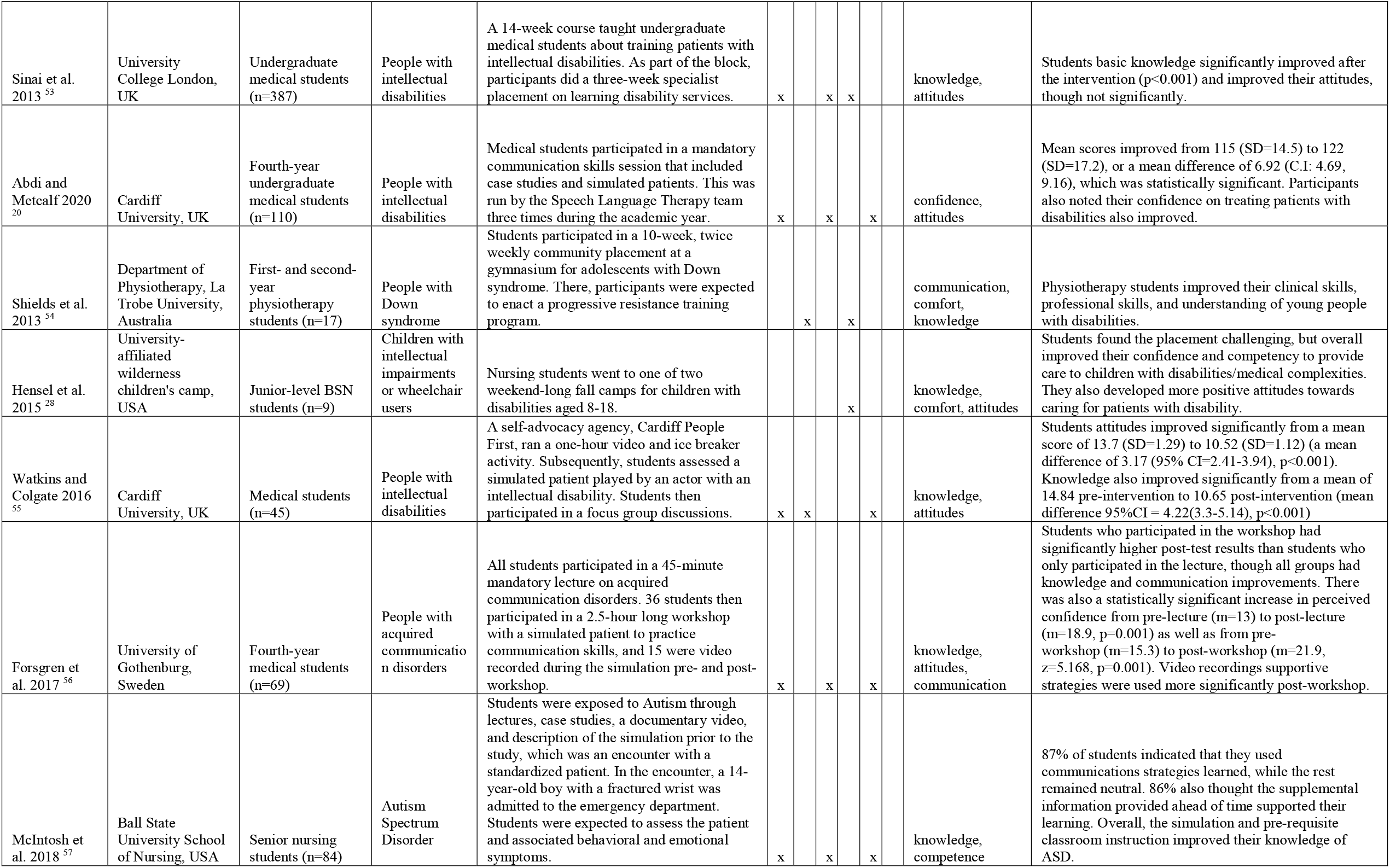
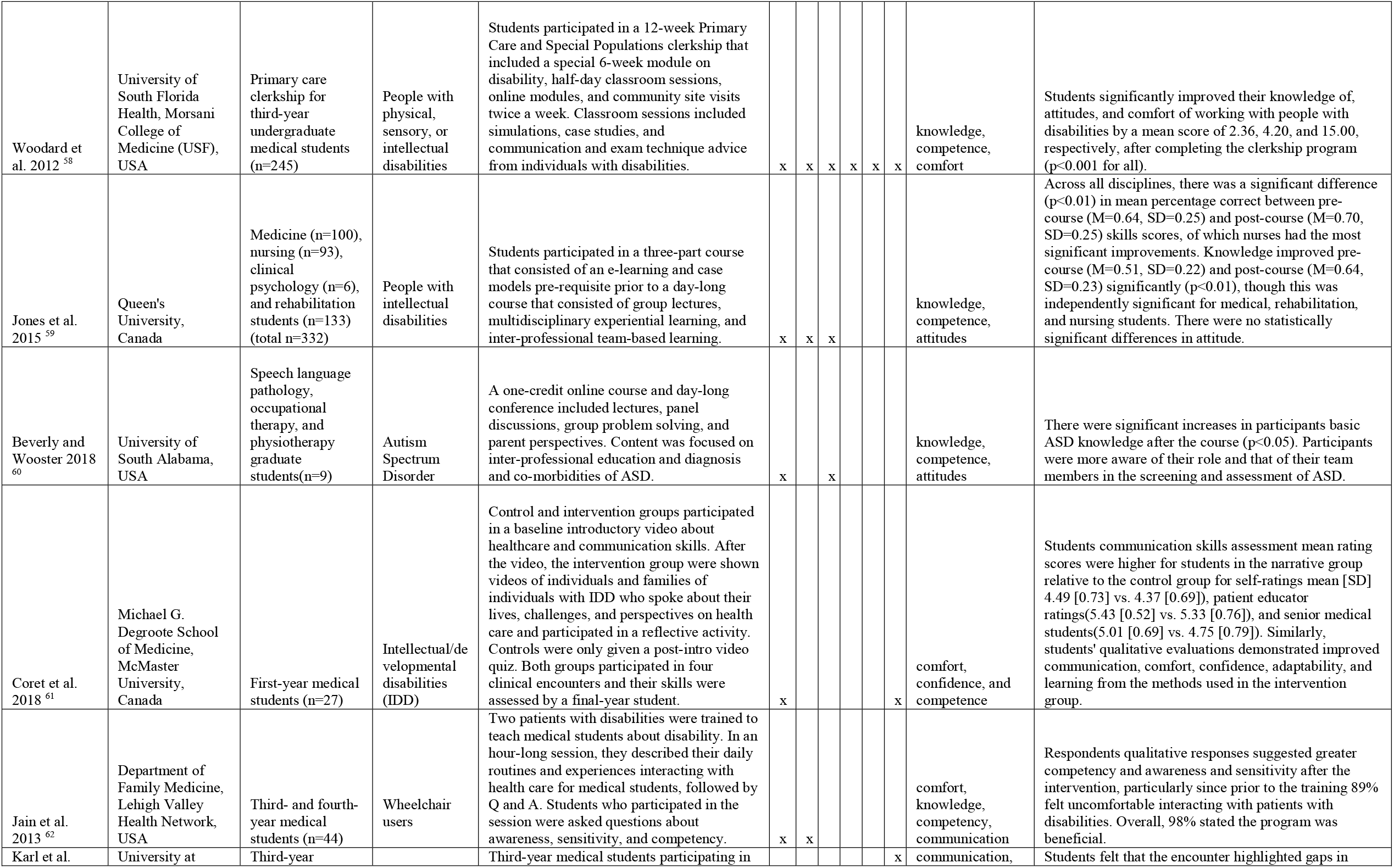
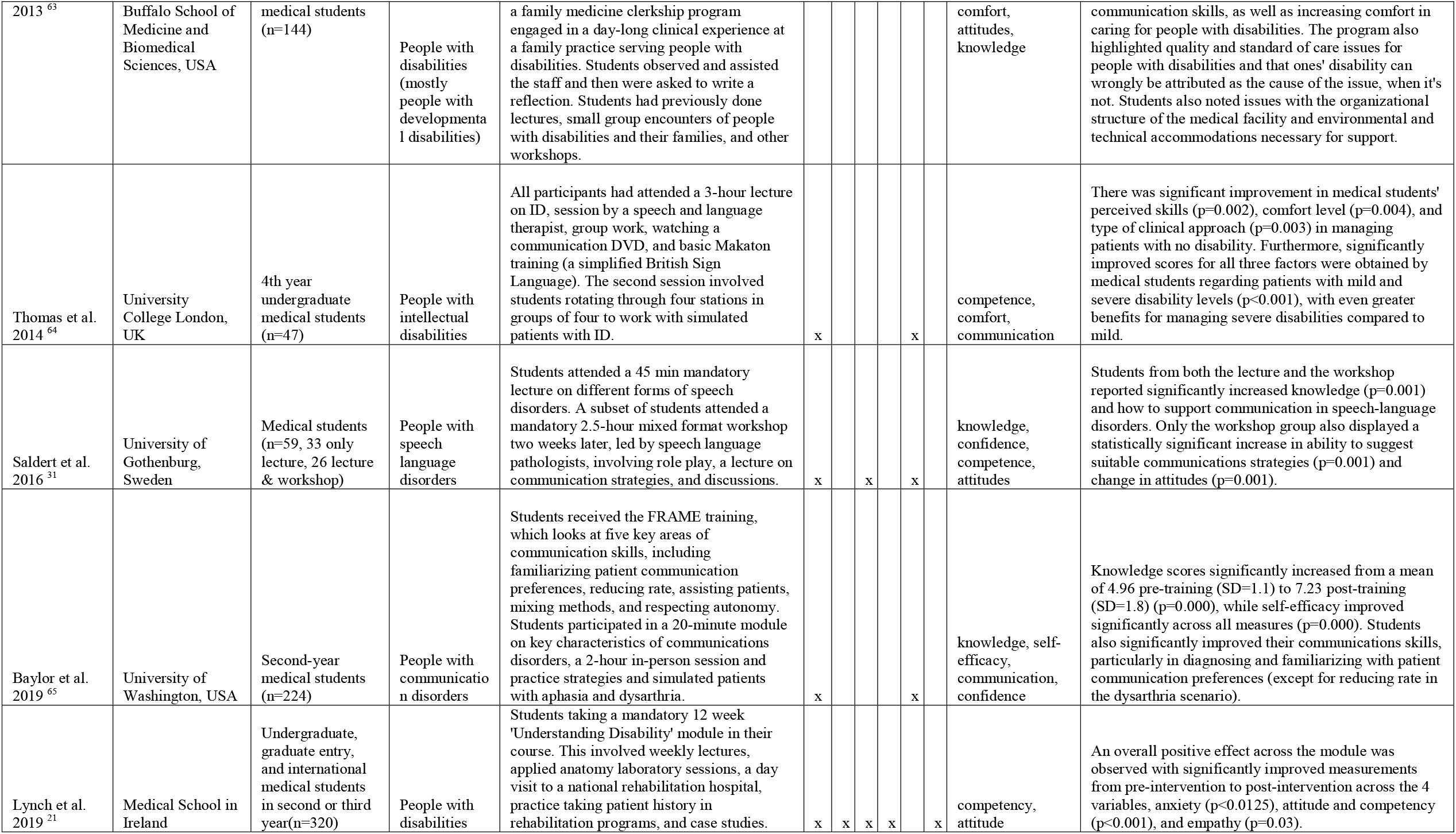
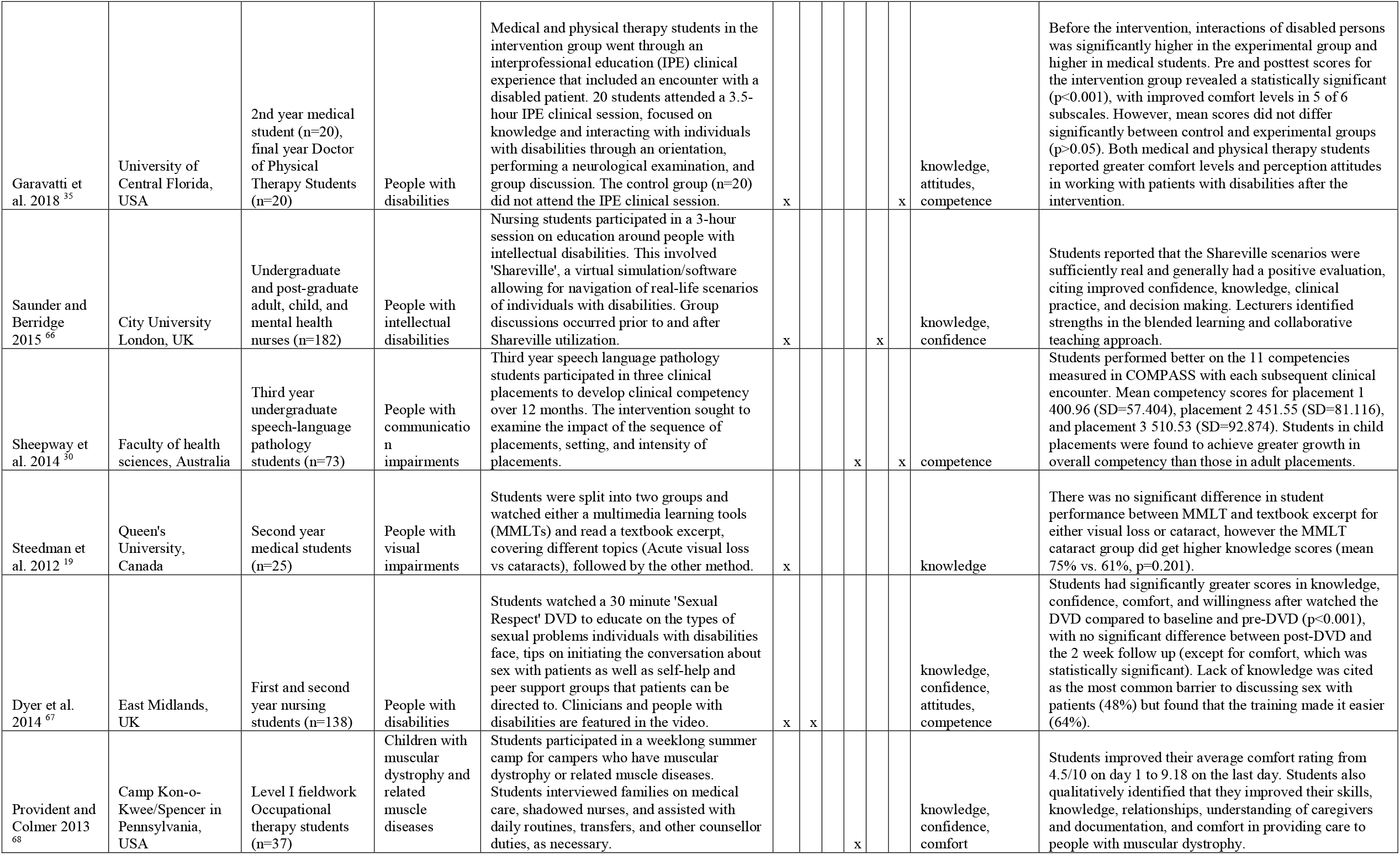
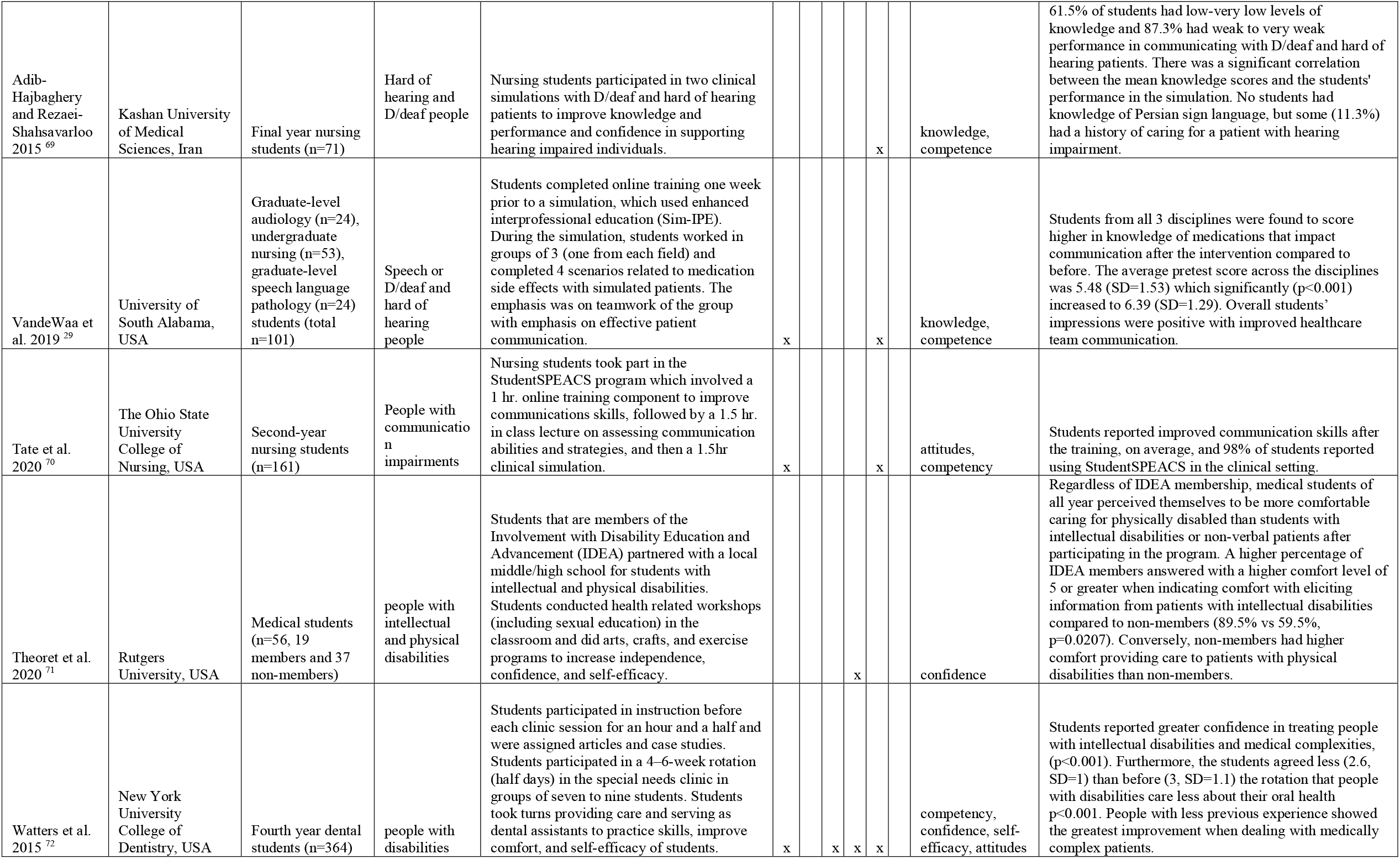
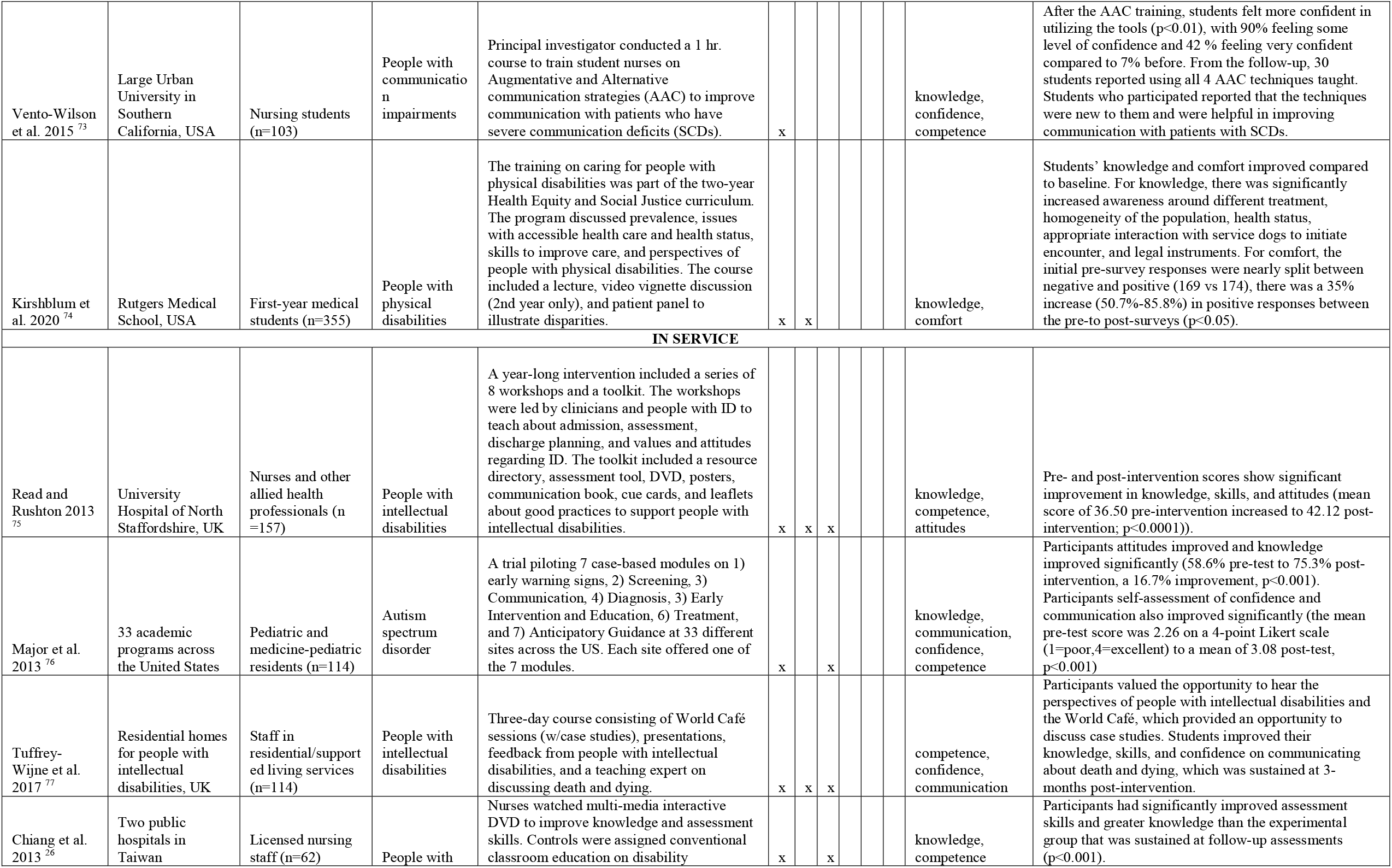
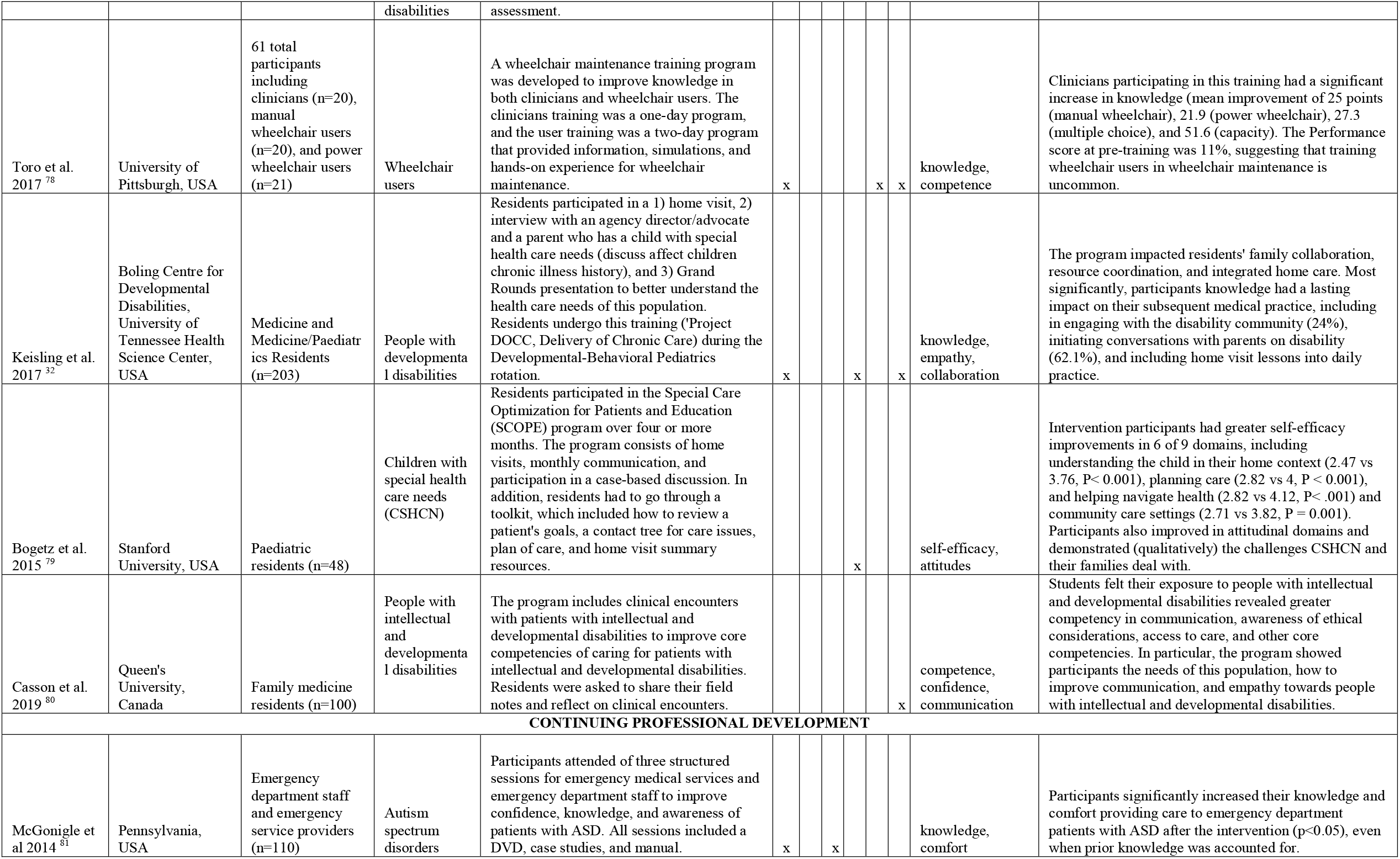
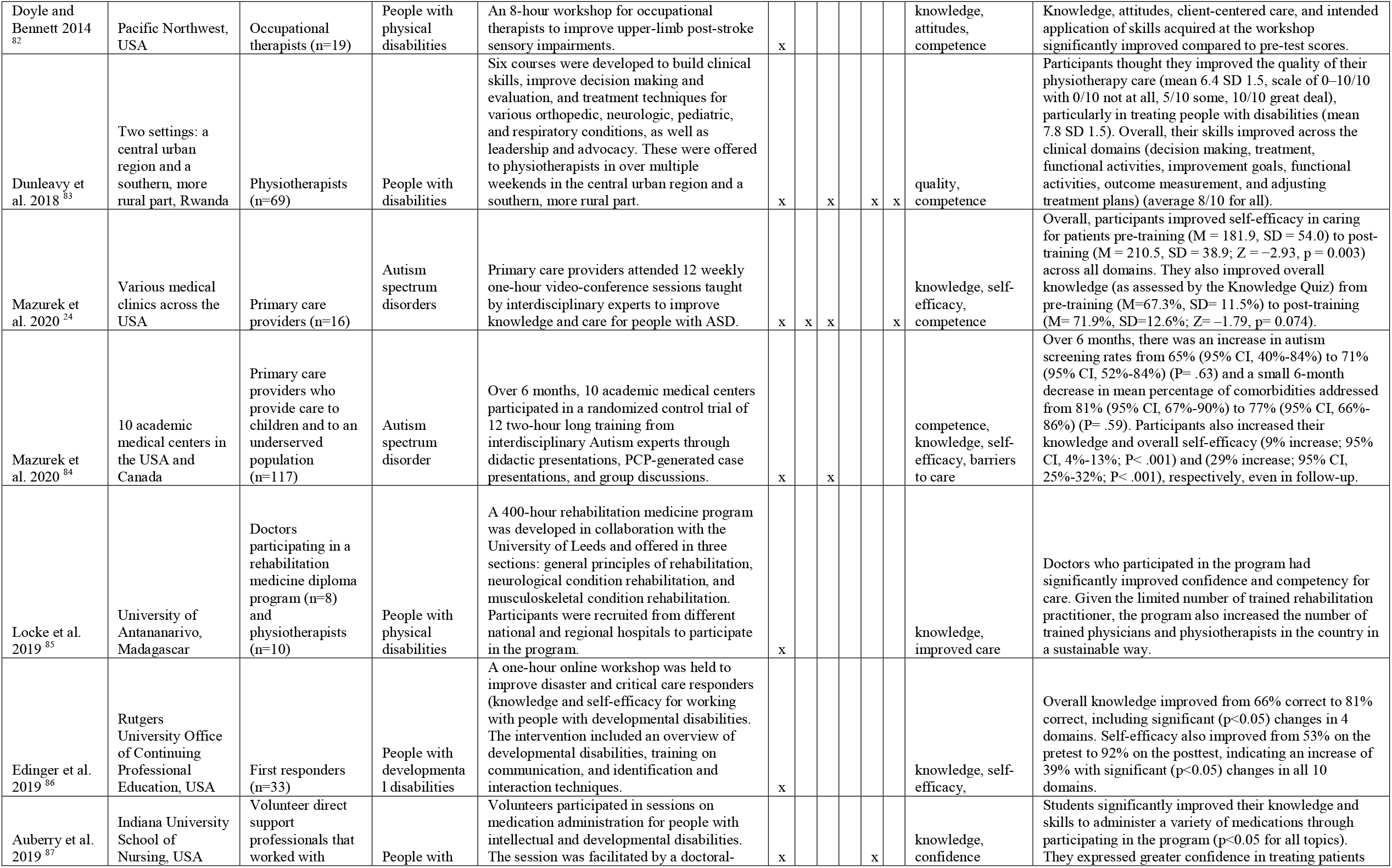
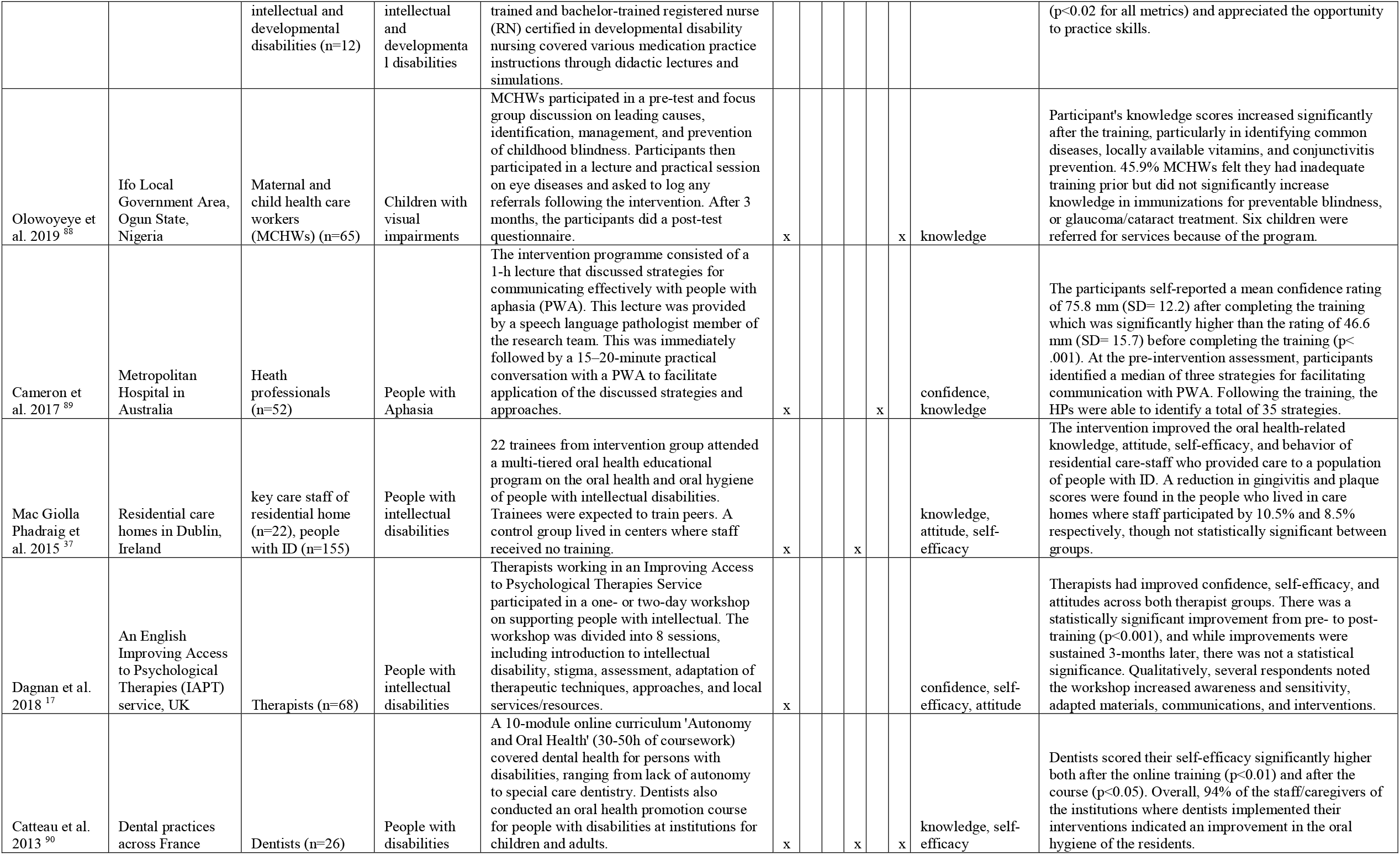
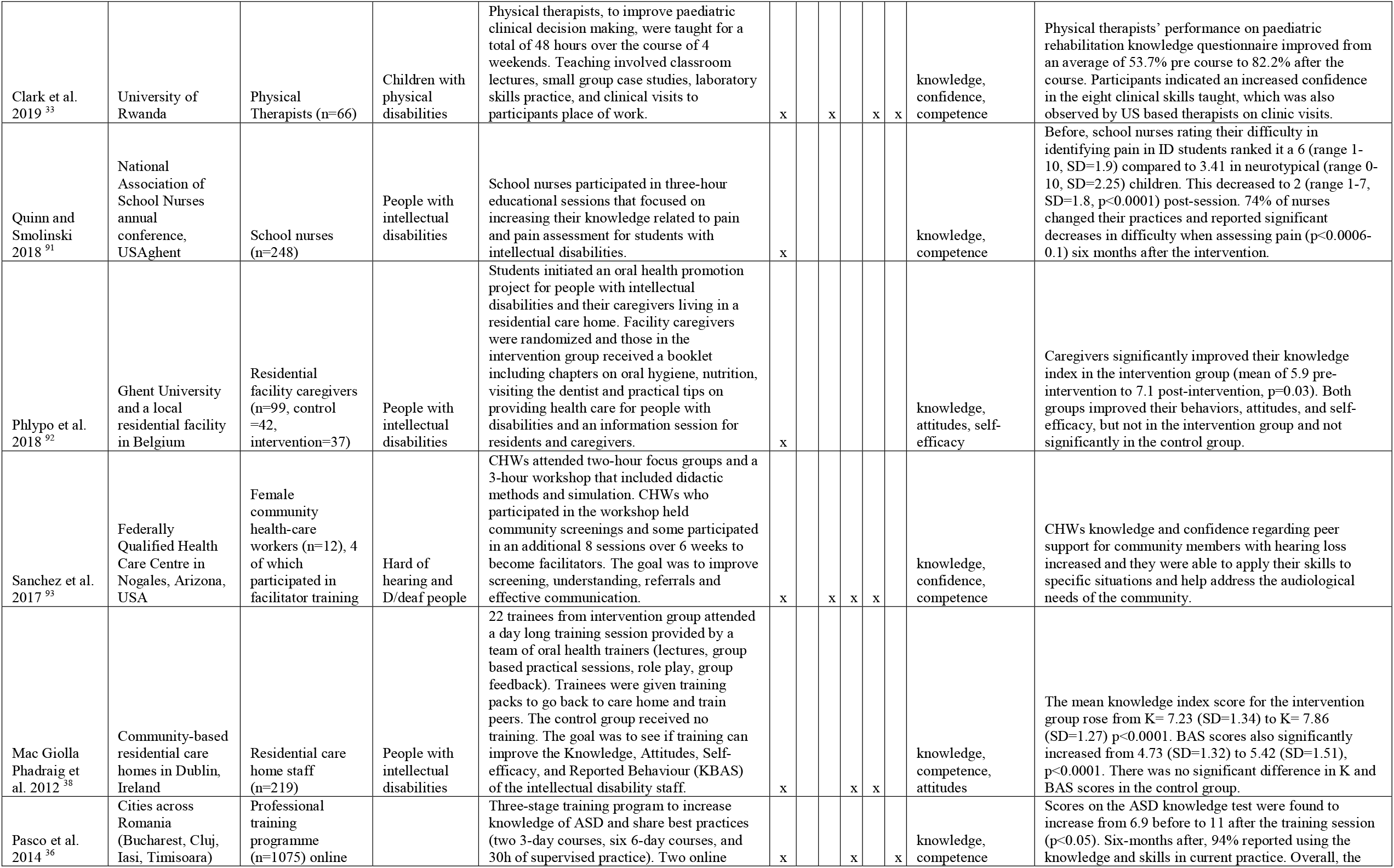
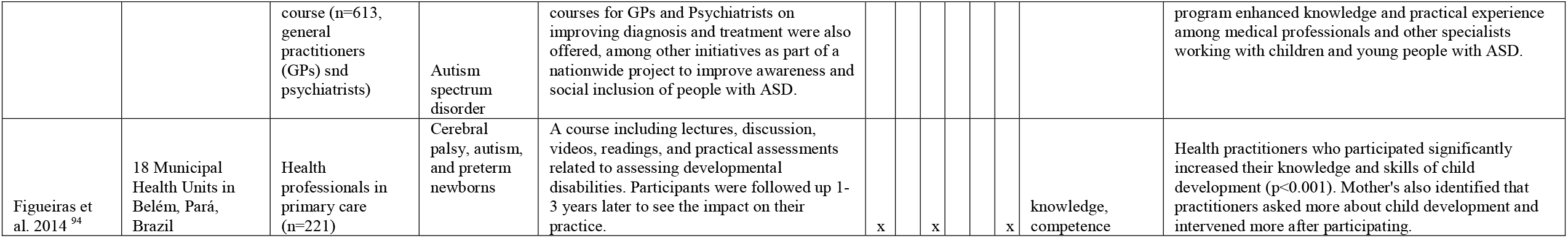

## Notes

### Competing Interest Statement

The authors have declared no competing interest.

### Funding Statement

This work was unfunded.

